# A tractometry investigation of white matter tract network structure and relationships with cognitive function in relapsing-remitting multiple sclerosis

**DOI:** 10.1101/2021.12.20.21268114

**Authors:** Danka Jandric, Geoff JM Parker, Hamied Haroon, Valentina Tomassini, Nils Muhlert, Ilona Lipp

## Abstract

Understanding the brain changes underlying cognitive dysfunction is a key priority in multiple sclerosis to improve monitoring and treatment of this debilitating symptom. Functional connectivity network changes are associated with cognitive dysfunction, but it is less well understood how changes in normal appearing white matter relate to cognitive symptoms. If white matter tracts share a similar network structure it would be expected that tracts within a network are similarly affected by MS pathology. In the present study, we used a tractometry approach to explore patterns of variance in diffusion metrics across white matter (WM) tracts. We investigated whether separate networks, based on normal variation or pathology, appear, and how this relates to neuropsychological test performance across cognitive domains. A sample of 102 relapsing-remitting MS patients and 27 healthy controls underwent MRI and neuropsychological testing. Tractography was performed on diffusion MRI data to extract 40 WM tracts and microstructural measures were extracted from each tract. Principal component analysis (PCA) was used to decompose metrics from all tracts to assess the presence of any co-variance structure among the tracts. Similarly, PCA was applied to cognitive test scores to identify the main cognitive domains. Finally, we assessed the ability of tract components to predict test performance across cognitive domains. We found that a single component which captured pathology across all tracts explained the most variance and that there was little evidence for separate, smaller network patterns of pathology. WM tract components were weak, but significant, predictors of cognitive function in MS. These findings highlight the need to investigate the relationship between the normal appearing white matter and cognitive impairment further and on a more granular level, to improve the understanding of the network structure of the brain in MS.

## 1. INTRODUCTION

The main pathological feature in multiple sclerosis is the demyelinating lesion, yet magnetic resonance imaging (MRI) measures of lesions correlate poorly with clinical symptoms, a finding termed the ‘clinico-radiological paradox’ (Barkhof, 1999, 2002). This is particularly problematic for cognitive symptoms, which affect a large proportion of people with MS, are disabling and associated with poor outcomes, but are poorly understood in terms of pathology (Sumowski *et al*., 2018), making cognitive dysfunction a challenge in the management of MS. Identifying MRI correlates of cognitive impairment in MS to understand pathological mechanisms is therefore a research priority to inform clinical decisions about diagnosis, monitoring and treatment of cognitive impairment, but a challenging one.

Cognitive deficits in MS are often evaluated as a global impairment based on results of neuropsychological tests. However, cognitive impairment involves deficits in separate domains, including processing speed and memory (Charcot, 1877; Benedict *et al*., 2006; Sepulcre *et al*., 2006; Migliore *et al*., 2016; Matias-Guiu *et al*., 2017; De Meo *et al*., 2021). Understanding if and how different cognitive domains are susceptible to different underlying brain abnormalities can inform our understanding of the mechanisms of cognitive impairment in MS.

Cognitive functions have been shown to rely on brain networks, rather than individual brain regions (McIntosh, 2000; Bressler and Menon, 2010). In people with MS, cognitive symptoms have been associated with functional network connectivity abnormalities, assessed by resting state fMRI (rs-fMRI, reviewed in Chard *et al*., 2021; Jandric *et al*., 2021), but the mechanisms causing these functional connectivity changes are not known. There is evidence to suggest that white matter (WM) damage can influence functional connectivity (Schoonheim *et al*., 2015; Patel *et al*., 2018; Tewarie *et al*., 2018), possibly through alteration of anatomical connections between functionally connected regions (Catani and ffytche, 2005; Dineen *et al*., 2009). While WM lesions are a poor predictor of cognitive symptoms, tissue outside of lesions is also known to be affected by pathological processes, either due to secondary axonal loss from inflammatory activity in lesions, such as Wallerian degeneration, or to lesion-independent degeneration of axons following demyelination resulting from more diffuse inflammation (Trapp *et al*., 1998; Bitsch *et al*., 2000; Trapp and Stys, 2009). Such damage to normal appearing white matter (NAWM) on a clinical MRI scan could cause damage to WM tracts connecting spatially separate but functionally connected regions that support specific cognitive functions. Moreover, if lesions cause damage to a brain network, the functioning of the entire network may be affected.

A number of diffusion MRI studies (dMRI) have established associations between cognitive impairment and damage to NAWM in MS. These studies have largely used whole-brain analyses of the WM, such as tract-based spatial statistics (TBSS, Smith *et al*., 2006), to show that non-lesional damage in specific WM areas, such as the corpus callosum and cingulum, correlates with cognitive symptoms (e.g. Dineen *et al*., 2009; Sbardella *et al*., 2013; Meijer *et al*., 2016). More recently, there has been evidence of covarying patterns of pathology in white matter tracts. In healthy participants, independent component analysis (ICA) based-approaches have demonstrated patterns of covariance between white matter tracts, thought to reflect shared phylogenetic and functional relationships (Wahl *et al*., 2010; Li *et al*., 2012). It can be expected that tracts that share characteristics and/or are part of the same networks are similarly susceptible to pathology. One study demonstrated this in a sample of secondary progressive MS patients (SPMS). Using ICA on a TBSS skeleton to identify patterns of covariance, possibly reflecting WM pathology, Meijer *et al*., (2016*a*) found eighteen components corresponding to WM tracts and visually grouped them into six different WM classes on the basis of anatomical features. FA values within some of these classes correlated with cognitive function, suggesting cognitively-relevant patterns of neurodegeneration (Meijer *et al*., 2016*a*).

Determining whether such patterns of pathology are also present in those with relapsing-remitting disease is necessary to identify at what stage in the disease such neurodegeneration occurs. While it is known that there is a greater degree of WM damage in SPMS (Kutzelnigg *et al*., 2005), people with RRMS also show both cognitive impairment and functional network changes, so it is plausible that they also show shared patterns of WM damage. It is also important to understand whether the patterns of WM damage appear when using an unbiased principal component analysis, which does not rely on manual grouping of component classes.

The standard for whole brain WM analysis has long been TBSS, which works by skeletonising the centre of each tract, based on high average FA values, to improve registration of non-homologous brains (Smith *et al*., 2006). As such TBSS does not reconstruct individual WM tracts, raising concerns about its anatomical accuracy (Bach *et al*., 2014). An alternative to TBSS for obtaining anatomically accurate WM tracts is tractography, which fits a diffusion tensor or alternative model at each voxel to trace the fibre orientation through the WM (Mori *et al*., 1999; Basser *et al*., 2000; Catani *et al*., 2002; Jeurissen *et al*., 2019). While challenging in its own right, technological developments have improved the ease and accuracy of individual, automated tratography (Warrington *et al*., 2020), and it has been shown that newer tracking algorithms can perform satisfactorily in the presence of MS lesions, and reconstruct even tracts with a high prevalence of lesions (Lipp *et al*., 2020). This makes tractography a feasible option for segmenting the brain into a large number of functionally meaningful WM units for investigating whether damage to non-lesioned parts of specific tracts can help understand cognitive symptoms in MS.

In the present study we conduct an exploratory analysis of WM microstructure diffusion metrics in a large sample of RRMS patients using a tractometry approach (Bells *et al*., 2011). We use automated individual tractography to reconstruct 40 WM tracts and extract four diffusion metrics from the non-lesioned parts of the tracts. By conducting principal component analysis (PCA) of extracted metrics we can test whether their grouping reflects the known network structure of the brain and covarying patterns of damage across tracts. Exploring this can help us understand the patterns of degeneration in normal appearing tissue in MS.

Thus, the present study aims to: 1) determine if WM tracts can be decomposed into components of shared covariance based on a network or pathology structure; 2) assess the cognitive domains structure present in common neuropsychological test data; 3) explore the relationship between WM tract components and cognitive domains in RRMS.

## 2. MATERIAL AND METHODS

### 2.1 Participants

Demographic, clinical and MRI data was collected in one study session from 102 RRMS patients and 27 healthy controls. This cohort has also been investigated and described in previous work (Jandric *et al*., 2021*b*). All participants were between 18 and 60 years of age, right-handed and had no contraindications for MR scanning. Patients fulfilled additional eligibility criteria of having no relapses or change to treatment for 3 months prior to the MRI scan, and not having any comorbid neurological or psychiatric disease.

Patients were recruited through the Helen Durham Centre for Neuroinflammation at the University Hospital of Wales and controls from the community. The study was approved by the NHS South-West Ethics and the Cardiff and Vale University Health Board R&D committees. All participants provided written informed consent to participate in the study.

### 2.2 Cognitive Assessment

Participants were assessed with the Multiple Sclerosis Functional Composite (MSFC) (Cutter *et al*., 1999) and the Brief Repeatable Battery of Neuropsychological Tests (BRB-N) (Amato *et al*., 2006). The BRB-N consists of the following tests: the selective reminding test of verbal memory, which is scored as the sum of words in long term storage (SRT L sum), the sum of words consistently recalled (SRT C sum) and the words recalled after a delay (SRT delayed); the spatial recall test of visual memory, which is scored over three consecutive trials (Spatial1to3) and on a delayed trial (Spatial delayed); the symbol digit modalities test (SDMT) of attention and concentration; the paced auditorial serial addition test of processing speed, with a three second delay (PASAT3) and with a two second delay (PASAT2); and the word list generation test (WLG) of verbal fluency.

### 2.3 MRI acquisition

MRI data were acquired on a 3 T MR scanner (General Electric HDx MRI System, GE Medical Devices, Milwaukee, WI) using an eight channel receive-only head RF coil. A high-resolution 3D T1-weighted sequence was acquired for identification of T1-hypointense MS lesions, segmentation, registration and volumetric measurements (voxel size = 1 mm x 1 mm x 1 mm, TE = 3.0 ms, TR = 7.8 ms, matrix = 256×256×172, FOV = 256 mm x 256 mm, flip angle = 20°). A T2/proton-density (PD)-weighted sequence (voxel size = 0.94 mm x 0.94 mm x 4.5 mm, TE = 9.0/80.6 ms, TR = 3000 ms, FOV = 240 mm x 240 mm, 36 slices, flip angle = 90°) and a fluid-attenuated inversion recovery (FLAIR) sequence (voxel size = 0.86 mm x 0.86 mm x 4.5 mm, TE = 122.3 ms, TR = 9502 ms, FOV = 220 mm x 220 mm, 36 slices, flip angle = 90°) were acquired for identification and segmentation of T2-hyperintense MS lesions. A twice refocused diffusion-weighted spin echo echo-planar (SE-EPI) sequence with 6 volumes with no diffusion weighting and 40 volumes with diffusion gradients applied in uniformly distributed directions was acquired for tractometrics analyses (diffusion directions: Camino 40, b = 1200 s/mm^2^, voxel size = 1.8 mm x 1.8 mm x 2.4 mm, TE = 94.5 ms, TR = 16000 ms, FOV = 230 mm x 230 mm, 57 slices, flip angle = 90°). In addition, a 3D MT sequence (voxel size = 0.94 mm x 0.94 mm x 1.9 mm, TE = 1.8 ms, TR = 26.7 ms, FOV = 240 mm x 240 mm, flip angle = 5°) and mcDESPOT sequence (voxel size = 1.7 mm x 1.7 mm x 1.7 mm, TE = SPGR: 2.1 ms, bSSFP: 1.6 ms, IR-SPGR: 2.1 ms, TR = SPGR: 4.7 ms, bSSFP: 3.2 ms, IR-SPGR: 4.7 ms, FOV = 220 mm x 220 mm, flip angle = SPGR: [3, 4, 5, 6, 7, 8, 9, 13, 18] degrees bSSFP: [10.6, 14.1, 18.5, 23.8, 29.1, 35.3, 45, 60] degrees IR-SPGR: 5 degrees) were acquired to obtain microstructure parameter maps as described in Lipp *et al*., (2019).

### 2.4 MRI processing

#### 2.4.1 Structural image analysis and lesion marking

Structural 3D T1-weighted images from patients were lesion filled, as described in previous work (Lipp *et al*., 2019), to allow better segmentation and registration of brain tissue, then segmented into grey matter (GM), WM and cerebrospinal fluid (CSF) using FSL’s Automated Segmentation Tool (FAST) (Zhang *et al*., 2001). Intracranial volume (ICV) was calculated with fslstats as the number of voxels in skull-stripped T1-weighted images. Volumetric measurements normalised for head size, including normalised brain volume (NBV), normalised GM volume (NGMV) and normalised WM volume (NAWM) were quantified from lesion-filled 3D T1-weighted images with FSL’s SIENAX tool (Smith *et al*., 2002). Lesion volume was calculated from binary lesion masks created as part of lesion marking. The lesion-filled 3D T1-weighted images were non-linearly registered to the Montreal Neurological Institute (MNI) 152 template space using FSL’s FNIRT tool and the warps saved for subsequent analyses.

#### 2.4.2 dMRI analysis: quantification of FA and RD maps

Preprocessing of dMRI data in ExploreDTI (v 4.8.3 (Leemans *et al*., 2009)) included motion correction and corrections for eddy current and EPI-induced geometrical distortions. The latter was achieved by registering each diffusion image to its respective (skull-stripped and downsampled to 1.5 mm) 3D T1 image (Irfanoglu *et al*., 2012) using Elastix (Klein *et al*., 2010), with appropriate reorientation of the diffusion-encoding vectors (Leemans and Jones, 2009). dMRI images were further processed in FSL’s FDT tool to fit diffusion tensors and fit the probabilistic diffusion model using the Bedpostx tool (Behrens *et al*., 2003, 2007). Fractional anisotropy (FA) and radial diffusivity (RD) maps were normalised to MNI space through the application of the previously obtained warps. FA and RD maps were available for all participants.

### 2.4. MTR and MWF maps

MTR and MWF maps were calculated as described in Lipp *et al*., (2019), which included co-registration with participants’ T1-weighted images. T1-weighted to MNI warps were applied for registration to MNI space. MTR maps were obtained for all HC and 101 MS patients, and MWF for 25 HC and 95 MS patients. MTR and MWF maps could not be obtained for some participants due to specific absorption rate (SAR) constraints of the mcDESPOT sequence, or due to logistical reasons.

### 2.5 Tractography and tractometry

Bedpostx outputs and T1-weighted to MNI registration warps were fed into FSL’s Xtract tool which uses standardised protocol seeding, exclusion, waypoint and termination masks to perform automated individual tractography to reconstruct 42 WM tracts, then uses the warps to register the outputs to MNI space (Warrington *et al*., 2020). All tracts were visually inspected to ensure that they had reconstructed well. In a large proportion of participants, both MS and HC, the fornix failed to reconstruct or was missing portions of the tract. As such, it was not considered for any analyses. The remaining 40 tracts yielded reconstructions in line with their anatomical descriptions and were retained.

Because the protocol masks are based on probability atlases of tracts, they are not strictly limited to the WM. To ensure that tract masks used for our analyses were limited to the WM to be suitable for tractometry analyses, we first thresholded the masked probabilistic tractography outputs at 0.001 and then masked the output further with the respective WM mask from the segmented T1 weighted scan and. These tracts were binarised and all voxels marked as lesions were removed to get a mask of only the non-lesioned part of the tract. The proportion of each tract affected by lesions in each participant was calculated by counting the lesion voxels in each tract relative to the voxels of the whole tract, averaged over all 102 participants for each tract.

To obtain FA, RD, MTR and MWF metrics from each reconstructed tract, each metric was averaged across all voxels in the non-lesioned tract masks. Distributions of each metric in each tract were assessed through histogram inspection in MATLAB (v R2020a). The majority of the FA, RD, MTR and MWF tract maps had distributions deviating from normality so median values were extracted.

#### 2.5.1 Metrics dimensionality reduction

The four WM microstructure metrics were extracted from each tract and decomposed into one metric using principal component analysis. This dimensionality reduction analysis was performed on the FA, RD, MWF and MTR metrics in RStudio v 1.4.1103 using the *principal* function (RStudio Team, 2020). Mean values were imputed for the missing MTR and MWF values and a dataset comprising of 4 WM metrics x 40 WM tracts x 27 or 102 participants (for HC and MS, respectively) was created. The four metrics were reduced to a lower dimensionality that explains the maximum amount of variance in the data through a PCA performed across participants and tracts, as described by Chamberland *et al*., (2019). First, a correlation matrix of Pearson’s r was calculated to determine feasibility of a PCA based on high correlations and tested with Bartlett’s test of sphericity to ensure a significant difference from an identity matrix. The metric principal component for further analyses was chosen on the basis of an inspection of the scree plot (Cattell, 1966) and eigenvalues >1. A metric component score for the first extracted principal component, explaining most variance, was calculated for each tract and participant.

#### 2.5.2 Principal component analysis of WM tract covariance

To assess whether patterns of shared covariance exist across the WM, an additional PCA, following the same process, was performed in HC and MS, respectively. For this PCA, the metric component score of the first extracted component was used as the WM microstructure metric for each tract.

#### 2.5.3 Regression of sources of heterogeneity in data

To identify the sources of variance in a tract component (TC) resulting from this PCA, its component scores were correlated with a number of demographic and anatomical variables: age, sex, years of education, ICV, lesion volume, NBV, NGMV and NWMV. Multivariate regressions were performed to identify which of these variables explained most variance of the TC. First, all demographic and anatomical were inputted into a correlation matrix to assess the degree of multicollinearity. As there was high correlation between NBV, NGMV and NWMV, only NBV was included in the model, along with age, sex, education ICV and lesion volume. The demographic and anatomical variables that came out as the strongest predictors in the regression analysis were regressed out of the raw data and the metric dimensionality reduction and PCA of WM tract covariance steps were performed again. The aim of this was to explore whether any potential heterogeneity in the sample which could have influenced the ability of the PCA to identify different components reliably. A Varimax rotation was applied to the first four principal components, based on eigenvalues >1 and proportion variance explained, to improve interpretability.

#### 2.5.4 Cognitive test principal component analysis

Finally, we aimed to find the cognitive domain structure in this dataset. As for the metric and tract PCAs, a correlation matrix was constructed based on the scores on each of the BRB-N tests, and on the basis of confirmed correlations between tests and a significant Bartlett’s test, a PCA was performed to decompose the battery tests into cognitive domains. Principal components were extracted on the basis of scree plots, eigenvalues and variance explained. A Varimax rotation was applied for interpretability. To understand what influences cognitive function, the resulting rotated cognitive components (CCs) were correlated with the tract components and all demographic and anatomical variables, after checking multicollinearity among predictors. NBV, NGMV and NWMV correlated highly so the variables included were age, sex, education, ICV, lesion volume and NBV. Multivariate regression analyses were performed to determine the relationship between WM tract microstructure and cognitive domains.

### 2.5 Statistical analyses

All analyses were performed in RStudio v 1.4.1103 (RStudio Team, 2020) with the exception of analyses of demographic and clinical variables, which were analysed in SPSS version 23.0 (IBM Corp., 2015). All variables were tested for normality through visual inspection of histograms and Q-Q plots and application of Kolmogorov-Smirnov tests. Variables which did not have a normal distribution were analysed with non-parametric tests.

## 3. RESULTS

### 3.1 Participant characteristics

Demographic and clinical characteristics of the sample are presented in Table 1. Overall, patients were older and less educated than healthy controls, had lower NBV and NGMV, poorer upper and lower limb function, and performed worse on all cognitive tests except the word list generation test assessing verbal fluency.

**Table 1.** Demographic, clinical and neuropsychological characteristics.

|  | **HC**<br>**(n=27)** | **RRMS**<br>**(n=102)** | *Inferential test results* |
| --- | --- | --- | --- |
| **Age, yr (median, range)** | 37.00 (23-59) | 45.00 (18-60) | $U = 958.00, p = .015$ |
| **Male/female, n** | 12/15 | 33/69 | $\chi^2(1) = 1.37, p = .241$ |
| **Education years (median, range)** | 19.00 (12-30) | 15.00 (10-30) | $U = 613.50, p < .001$ |
| **Mean disease duration, yr (median, range)** | N/A | 12.24 (1-39) | N/A |
| **Timed 25 Foot Walk Test (median, range)** | 4.35 (3.2-5.4) | 5.25 (3.6-26.8) | $U = 572.50, p < .001$ |
| **9-Hole Peg Test (median, range)** | 18.65 (15.35-23.00) | 21.75 (16.35-59.50) | $U = 537.50, p < .001$ |
| **SRT L sum (median, range)** | 0.00 (-1.26-1.37) | -0.54 (-4.72-1.47) | $U = 914.00, p = .009$ |
| **SRT C sum (mean, SD)** | 0.00 (1.00) | -0.88 (1.22) | $t(49.06) = 3.86, p < .001$ |
| **SRT delayed (median, range)** | 0.06 (-2.13-1.16) | -0.49 (-4.31-1.15) | $U = 881.00, p = .004$ |
| **Spatial1to3** | 0.00 (1.00) | -0.74 (1.20) | $t(47.76) = 3.29, p = .002$ |
| **Spatial delayed** | 0.11 (-2.45-1.14) | -0.91 (-2.96-1.14) | $U = 794.00, p = .001$ |
| **SDMT** | 0.00 (1.00) | -0.88 (0.98) | $t(40.14) = 4.09, p < .001$ |
| **PASAT3 (median, range)** | -0.03 (-2.61-1.26) | -1.32 (-8.26-1.42) | $U = 692.00, p < .001$ |
| **PASAT2** | 0.17 (-1.71-2.29) | -0.77 (-4.45-2.41) | $U = 768.00, p < .001$ |
| **WLG** | 0.00 (1.00) | -0.24 (0.88) | $t(37.48) = 1.14, p = .263$ |
| **Normalised brain volume, L (mean, SD)** | 1.56 (0.07) | 1.51 (0.08) | $t(41.94) = 3.33, p = .002$ |
| **Normalised grey matter volume, L (median, range)** | 0.81 (0.72-0.89) | 0.77 (0.61-0.89) | $U = 755.00, p < .001$ |
| **Normalised white matter volume, L (median, range)** | 0.76 (0.68- 0.81) | 0.74 (0.66- 0.83) | $t(40.43) = 1.56, p = .127$ |

Independent samples t-tests were used for comparisons of variables with a normal distribution. Mann-Whitney U tests were used for variables which were not normally distributed. Sex, being a categorical variable, was tested with the chi-squared test. Cognitive test scores are reported as Z-scores.
Abbreviations: HC = healthy controls; PASAT2 = paced auditory serial addition test 2 second delay; PASAT3 = paced auditory serial addition test 3 second delay; RRMS = relapsing remitting multiple sclerosis; SD = standard deviation; SDMT = symbol digit modalities test; Spatial1to3 = spatial recall test average score over three trials; Spatial delayed = spatial recall test score at the delayed trial; SRT delayed = serial recall test scores at the delayed trial; SRT L sum = serial recall test long term storage sum of scores; SRT L sum = serial recall test consistent recall sum of scores; WLG = word list generation test.

### 3.2 Metric dimensionality reduction

The following results are for the MS group unless otherwise indicated. Detailed results for healthy controls are presented in Appendix 1.

Bartlett’s test of sphericity was significant (χ^2^(6) = 155.85, *p* <0.001) indicating the suitability of performing a PCA. Based on scree plot inspection and eigenvalues >1, only the first principal component, which explained 60% of variance, was extracted. The component loadings were 0.92 for FA, -0.87 for RD, 0.88 for MWF and 0.15 for MTR, indicating that the main contributors to the component were FA, RD and MWF.

### 3.3 Principal components of WM tract covariance

In MS patients, a correlation matrix of all WM tracts was shown to be significantly different from an identity matrix using Bartlett’s test of sphericity (χ^2^(780) = 5803.14, *p* <0.001), indicating the suitability of performing a PCA to assess the covariance structure of WM tracts (see Figure 1A for the metric and tract correlation matrices and scree plots). The scree plot showed one strong principal component (65% variance explained), but three additional components had eigenvalues >1 (7%, 4% and 3% variance explained, respectively). A Varimax rotation was therefore applied to these first four principal components to improve interpretability. After rotation all tracts still loaded positively on TC1, demonstrating a great degree of shared variable between white matter tracts.

**Figure 1.**
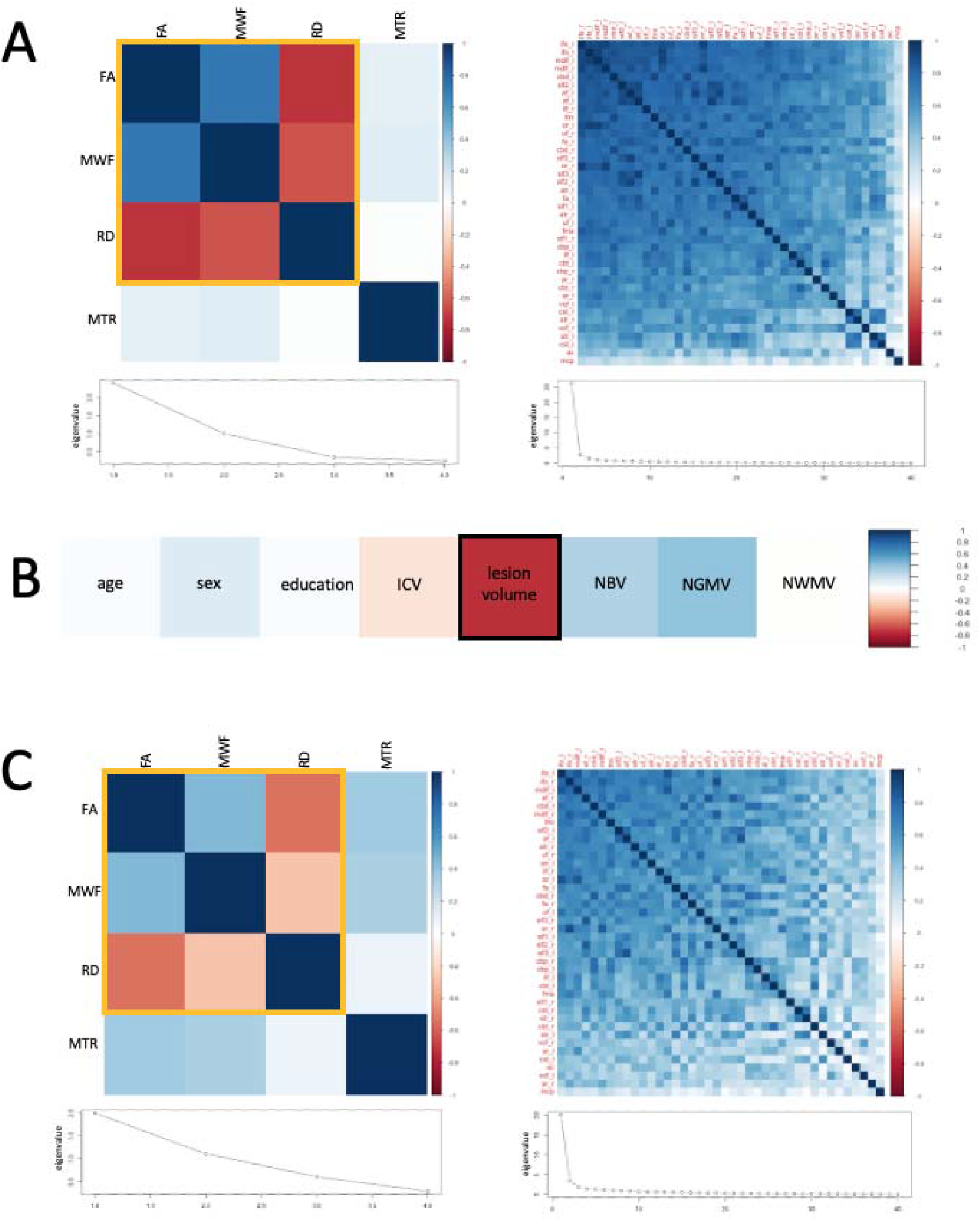
Metric and tract principal component analysis in healthy controls. Figure 1A shows the correlation matrices and scree plots for the PCA ran on the four white matter microstructural metrics (left) and the white matter tracts based on the first component from the metric PCA (right). Those metrics marked with a yellow line load most on principal component 1. All tracts loaded positively on tract principal component 1. Figure 1B shows correlations between rotated tract principal component 1 (TC1) and demographic and anatomical variables. Variable marked with a black line, lesion volume, was a significant predictor of the principal tract component from Figure 1A in multiple linear regression models. Figure 1C shows the correlation matrices and scree plots for metric and tract PCAs after lesion volume was regressed out. Abbreviations: FA = fractional anisotropy; RD = radial diffusivity; MWF = myelin water fraction; MTR = magnetisation transfer ratio; ICV = intracranial volume; NBV = normalised brain volume; NGMV = normalised grey matter volume; NWMV = normalised white matter volume

In MS patients TC1 correlated most strongly with lesion volume (r=-0.73), NGMV (r=0.41), and NBV (r=0.31) (see Figure 1B). A multiple linear regression model showed that the variance of TC1 was best explained by lesion volume (ß = -0.74, *p* < 0.001) in a model explaining 54% of variance (R^2^ = 0.54, F(6, 95) = 20.58, *p* < 0.001). See Table 2 for full model statistics.

**Table 2.** Predictors of WM tract covariance and cognitive domains.

| **Dependent variable** | **Predictors** | **Model statistics** |
| --- | --- | --- |
| **Unrotated tract component 1** | Age: $\beta = 0.03$, $p = 0.727$<br>Sex: $\beta = 0.13$, $p = 0.186$<br>Education: $\beta = -0.03$, $p = 0.690$<br>ICV: $\beta = -0.03$, $p = 0.721$<br>*Lesion volume: $\beta = -0.74$, $p < 0.001$*<br>NBV: $\beta = 0.01$, $p = 0.451989$ | $R^2 = 0.54$, $F(6, 95) = 20.58$, $p < 0.001$ |
| **CC1: Verbal cognition** | *TC1: $\beta = 0.30$, $p = 0.009$*<br>TC2: $\beta = -0.07$, $p = 0.444$<br>TC3: $\beta = 0.14$, $p = 0.120$<br>TC4: $\beta = 0.08$, $p = 0.379$<br>Age: $\beta = -0.01$, $p = 0.957$<br>*Sex: $\beta = 0.35$, $p = 0.010$*<br>Education: $\beta = 0.06$, $p = 0.558$<br>ICV: $\beta = 0.12$, $p = 0.365$<br>*Lesion volume: $\beta = -0.22$, $p = 0.049$*<br>*NBV: $\beta = -0.32$, $p = 0.028$* | $R^2 = 0.17$, $F(10, 91) = 3.04$, $p < 0.001$ |
| **CC2: Visuospatial cognition** | TC1: $\beta = -0.01$, $p = 0.938$<br>TC2: $\beta = -0.13$, $p = 0.191$<br>TC3: $\beta = 0.08$, $p = 0.442$<br>TC4: $\beta = -0.11$, $p = 0.290$<br>*Age: $\beta = -0.32$, $p = 0.009$*<br>Sex: $\beta = 0.10$, $p = 0.482$<br>Education: $\beta = 0.09$, $p = 0.358$<br>ICV: $\beta = 0.01$, $p = 0.963$<br>Lesion volume: $\beta = -0.04$, $p = 0.708$<br>NBV: $\beta = -0.04$, $p = 0.800$ | $R^2 = 0.05$, $F(10, 91) = 1.50$, $p = 0.153$ |
| **CC3: Information processing** | TC1: $\beta = 0.06$, $p = 0.628$<br>TC2: $\beta = 0.12$, $p = 0.236$<br>*TC3: $\beta = 0.24$, $p = 0.017$*<br>TC4: $\beta = -0.16$, $p = 0.101$<br>Age: $\beta = 0.18$, $p = 0.127$<br>Sex: $\beta = -0.14$, $p = 0.318$<br>Education: $\beta = -0.02$, $p = 0.839$<br>ICV: $\beta = -0.10$, $p = 0.493$<br>Lesion volume: $\beta = 0.04$, $p = 0.739$<br>NBV: $\beta = 0.20$, $p = 0.197$ | $R^2 = 0.06$, $F(10, 91) = 1.65$, $p = 0.106$ |
| **CC4: Executive function** | TC1: $\beta = -0.02$, $p = 0.870$<br>TC2: $\beta = 0.05$, $p = 0.604$<br>TC3: $\beta = 0.15$, $p = 0.101$<br>TC4: $\beta = 0.03$, $p = 0.747$<br>*Age: $\beta = -0.27$, $p = 0.016$*<br>Sex: $\beta = -0.03$, $p = 0.851$<br>Education: $\beta = 0.07$, $p = 0.443$<br>ICV: $\beta = -0.02$, $p = 0.872$ | $R^2 = 0.18$, $F(10, 91) = 3.19$, $p = 0.002$ |
| | *Lesion volume: $\beta = -0.23$, $p = 0.035$*<br>NBV: $\beta = 0.13$, $p = 0.352$ | |

Significant predictors are presented in italics. Significance threshold $p < 0.05$ applied unless otherwise indicated. Abbreviations: CC = cognitive component , NBV = normalised brain volume, NWMV = normalised white matter volume, TC = tract component, WM = white matter

After regressing out lesion volume, correlations matrices for WM metrics and tracts, respectively, showed somewhat weaker correlations but still passed Bartlett’s test of sphericity (χ^2^(6) = 90.01, *p* < 0.001 for metrics, χ^2^(780) = 4347.86, *p* < 0.001 for tracts), and yielded the same PCA structure (see Figure 1C), indicating that most tracts still load positively onto a single component. After a component rotation of the four tracts that explained most of the variance (after rotation:79% cumulative variance; 30%, 25%, 21%, 0.03% for TCs 1-4, respectively) most tracts still loaded positively on the first tract component, especially large WM tracts like the optic radiations, middle longitudinal fasciuli, forceps major, inferior fronto-occipital fasciculi, vertical occipital fasciculi and acoustic radiations. Similarly, the tracts which loaded most highly on TC2 were large tracts connecting distal areas of the brain, including the superior thalamic radiations, corticospinal tracts, frontal aslants, superior longitudinal fasiculi and the arcuate fasciculi. TC3 in contrast consisted mainly of shorter tracts, including sub-sections of the cingulum, the anterior commissure, forceps minor and uncinate fasciuli. Only the middle cerebellar penduncle loaded highly on TC4. The principal component analysis screeplot showing a single dominant component and the high tract loadings of all tracts onto the first of the four rotated components demonstrates a high covariance between all tracts investigated. Please see Table 3 for full details of tract loadings on the four components.

**Table 3.** Tract loadings on each component derived from the tract PCA, after regressing out significant predictors of tract variance and applying Varimax rotation.

| TC1 |  | TC2 |  | TC3 |  | TC4 |  |
| --- | --- | --- | --- | --- | --- | --- | --- |
| Tract | Loading | Tract | Loading | Tract | Loading | Tract | Loading |
| or_l | 0.77 | str_l | 0.89 | cbp_r | 0.77 | mcp | 0.84 |
| mdlf_l | 0.75 | cst_l | 0.86 | ac | 0.73 | vof_r | 0.26 |
| or_r | 0.75 | str_r | 0.82 | cbd_r | 0.72 | ac | 0.25 |
| fma | 0.75 | cst_r | 0.78 | cbt_r | 0.70 | fma | 0.23 |
| mdlf_r | 0.74 | fa_l | 0.73 | cbt_l | 0.67 | ar_r | 0.23 |
| ifo_r | 0.72 | fa_r | 0.68 | cbp_l | 0.67 | or_r | 0.23 |
| vof_l | 0.70 | af_l | 0.66 | fmi | 0.65 | cst_r | 0.23 |
| ar_l | 0.70 | slf1_l | 0.65 | uf_l | 0.64 | atr_r | 0.21 |
| vof_r | 0.69 | slf3_l | 0.65 | cbd_l | 0.63 | atr_l | 0.18 |
| ar_r | 0.68 | af_r | 0.63 | uf_r | 0.60 | mdlf_r | 0.16 |
| ifo_l | 0.67 | slf2_r | 0.62 | atr_l | 0.59 | ifo_r | 0.14 |
| ilf_r | 0.67 | slf2_l | 0.59 | atr_r | 0.58 | ilf_l | 0.12 |
| slf2_l | 0.62 | atr_l | 0.59 | ilf_l | 0.55 | fmi | 0.12 |
| af_r | 0.62 | slf3_r | 0.59 | ifo_l | 0.50 | af_r | 0.09 |
| slf1_r | 0.61 | atr_r | 0.56 | ilf_r | 0.46 | or_l | 0.09 |
| slf3_r | 0.60 | mdlf_r | 0.47 | mdlf_l | 0.44 | slf3_r | 0.08 |
| ilf_l | 0.58 | slf1_r | 0.44 | ifo_r | 0.43 | cbt_r | 0.08 |
| slf2_r | 0.58 | ifo_l | 0.44 | fa_l | 0.39 | ilf_r | 0.07 |
| af_l | 0.56 | ifo_r | 0.43 | slf1_l | 0.39 | str_r | 0.06 |
| cbd_l | 0.56 | cbd_l | 0.42 | fma | 0.38 | uf_r | 0.04 |
| uf_r | 0.55 | fmi | 0.41 | slf2_l | 0.37 | slf2_r | 0.03 |
| cbt_l | 0.54 | mdlf_l | 0.40 | or_l | 0.37 | ifo_l | 0.02 |
| slf3_l | 0.53 | or_r | 0.40 | ar_l | 0.37 | vof_l | 0.02 |
| fa_r | 0.52 | ilf_r | 0.39 | af_l | 0.34 | str_l | 0.02 |
| fmi | 0.49 | uf_r | 0.37 | mdlf_r | 0.33 | cbd_r | 0.02 |
| cbd_r | 0.47 | uf_l | 0.37 | fa_r | 0.33 | cbd_l | 0.01 |
| uf_l | 0.45 | cbd_r | 0.35 | vof_l | 0.33 | cbt_l | 0.00 |
| slf1_l | 0.44 | or_l | 0.35 | slf1_r | 0.32 | mdlf_l | 0.00 |
| cbt_r | 0.44 | ar_r | 0.32 | slf3_l | 0.32 | slf3_l | -0.03 |
| cbp_l | 0.42 | cbp_l | 0.31 | af_r | 0.31 | cst_l | -0.03 |
| fa_l | 0.38 | cbp_r | 0.27 | slf3_r | 0.30 | slf1_r | -0.03 |
| atr_r | 0.31 | ilf_l | 0.21 | or_r | 0.29 | af_l | -0.04 |
| atr_l | 0.31 | fma | 0.21 | slf2_r | 0.28 | cbp_l | -0.04 |
| cbp_r | 0.31 | vof_r | 0.15 | vof_r | 0.21 | fa_r | -0.04 |
| cst_r | 0.24 | ar_l | 0.14 | ar_r | 0.17 | slf2_l | -0.05 |
| mcp | 0.22 | cbt_l | 0.13 | str_r | 0.16 | cbp_r | -0.05 |
| str_r | 0.19 | vof_l | 0.11 | cst_r | 0.14 | slf1_l | -0.06 |
| str_l | 0.14 | cbt_r | 0.11 | mcp | 0.13 | uf_l | -0.07 |
| cst_l | 0.08 | ac | 0.04 | cst_l | 0.09 | fa_l | -0.12 |

Abbreviations: ac = anterior commissure; af = arcuate fasciculus; ar = acoustic radiation; atr = anterior thalamic radiation; cbd = cingulum subsection, dorsal; cbp = cingulum subsection, peri-genua; cbt = cingulum
subsection, temporal; cst = corticospinal tract; fa = frontal aslant; fma = forceps major; fmi = forceps minor; ifo = inferior fronto-occipital fasciculus; ilf = inferior longitudinal fasciculus; mcp = middle cerebellar peduncle; mdlf = middle longitudinal fasciculus; or = optic radiation; slf1-3 = superior longitudinal fasciculus 1-3; str = superior thalamic radiation; uf = uncinate fasciculus; vof = vertical occipital fasciculus. Left and right hemisphere tracts are denoted with \_l and \_r, respectively.

### 3.4 Cognitive domains

A correlation matrix of cognitive test scores showed a large number of moderate to high correlations and was significantly different from an identity matrix as assessed by Bartlett’s test of sphericity (χ^2^(36) = 558.62, *p* <0.001), indicating a likely domain structure of cognition and confirming suitability for a PCA. Based on eigenvalues of at or near 1 and proportion variance explained, four components explaining 85% of variance were extracted. After a Varimax rotation a clear component structure emerged whereby cognitive component (CC) 1 reflects verbal cognition and CC2 visuospatial cognition, while CCs 3 and 4 reflect information processing speed and executive function, respectively. The component weights for rotated cognitive components (CCs) were as follows: Serial Recall Test Consistent recall (0.87), Serial Recall Test Long term storage recall (0.82), Word List Generation Test (0.80) and Serial Recall Test delayed recall (0.73) for CC1; Spatial Recall Test over three trials (0.90) and Spatial Recall Test delayed recall (0.93) for CC2; Paced Auditory Serial Addition Test 3 second delay (0.88) and Paced Auditory Serial Addition Test 2 second delay (0.89) for CC3; and Symbol Digit Modalities Test (0.84) for CC4, see Table 4.

**Table 4.** Cognitive component weights in MS patients.

|  | **Cognitive RC1** | **Cognitive RC2** | **Cognitive RC3** | **Cognitive RC4** |
| --- | --- | --- | --- | --- |
| **SRT L sum** | **0.82** | 0.16 | 0.10 | 0.36 |
| **SRT C sum** | **0.87** | 0.18 | 0.21 | 0.24 |
| **SRT delayed** | **0.73** | 0.25 | 0.29 | 0.37 |
| **Spatial1to3** | 0.15 | **0.90** | 0.23 | 0.06 |
| **Spatial delayed** | 0.10 | **0.93** | 0.01 | 0.15 |
| **SDMT** | 0.20 | 0.17 | 0.27 | **0.84** |
| **PASAT3** | 0.17 | 0.18 | **0.88** | 0.20 |
| **PASAT2** | 0.25 | -0.06 | **0.89** | 0.11 |
| **WLG** | **0.80** | -0.05 | 0.17 | -0.29 |

Abbreviations: PASAT2 = paced auditory serial addition test 2 second delay; PASAT3 = paced auditory serial addition test 3 second delay; SDMT = symbol digit modalities test; Spatial1to3 = spatial recall test average score over three trials; Spatial delayed = spatial recall test score at the delayed trial; SRT delayed = serial recall test scores at the delayed trial; SRT L sum = serial recall test long term storage sum of scores; SRT C sum = serial recall test consistent recall sum of scores; WLG = word list generation test.

### 3.5 Tract components are weak predictors of cognition

In MS, tract components were modest to weak predictors of cognitive components, as were demographic and MRI variables (see Table 2). The first cognitive component, CC1, was best predicted by TC1 (ß = 0.30, p = 0.009), sex (ß = 0.35, p = 0.010), lesion volume (ß = -0.22, p = 0.049) and NBV (ß = -0.32, p = 0.028), in a model explaining 17% of variance (R^2^ = 0.17, F(10, 91) = 3.04, p < 0.001). The final cognitive component, CC4, was best predicted by age (ß = -0.27, p = 0.016) and lesion volume (ß = -0.23, p = 0.035), in a model explaining 18% of the variance (R^2^ = 0.18, F(10, 91) = 3.19, p = 0.002). For cognitive components 2 and 3, the regression models were not significant. Given the low predictive values of tract components on cognitive components, WM variance patterns are weakly linked to cognitive domains. Please see Table 2 for full statistical results.

## 4. DISCUSSION

In this study we combined PCA with tractometry to determine whether cognitive performance in people with MS relates to one or many patterns of white matter tract pathology. A decomposition approach of microstructure metrics from WM tracts showed a high degree of covariance across most tracts, indicating a global WM structure rather than a network-specific structure. This global WM microstructure component was largely explained by lesion volume, but retained largely a single covariance pattern even after this factor was regressed out. Cognitive domains were only weakly explained by WM microstructure components. This demonstrates that changes in white matter microstructure in people with MS is dominated by a single pattern of pathology, which is weakly associated with impaired cognition.

### 4.1 Metric dimensionality reduction

Tract decomposition was based on several diffusion metrics combined into one, consisting of FA, RD and MWF. Traditionally FA is used in MS studies of cognition, but FA has been shown to be susceptible to many factors, including myelination, axonal density and orientational dispersion of fibre populations in a voxel (Beaulieu, 2014; De Santis *et al*., 2014; Lazari and Lipp, 2021). A multimodal approach is a useful alternative for obtaining more comprehensive information about WM microstructure, and has been shown to produce a sensitive component measure of WM microstructure when several metrics are combined in a tractometry approach (Chamberland *et al*., 2019; Geeraert *et al*., 2020; Bosticardo *et al*., 2021).

Such dimensionality reduction has been shown to overcome the problem of multiple comparisons of data containing overlapping information while maintaining good sensitivity of WM microstructure (Chamberland *et al*., 2019; Geeraert *et al*., 2020; Bosticardo *et al*., 2021). Recently is has been shown to provide a more sensitive measure of MS pathology for connectomics approaches than the number of streamlines traditionally used (Bosticardo *et al*., 2021).

### 4.2 WM microstructure organisation

A number of studies have demonstrated functional network changes (Chard *et al*., 2021, Jandric *et al*., 2021*a*) and patterns of grey and white matter pathology associated with cognitive impairment in MS (Meijer *et al*., 2016*a*; Steenwijk *et al*., 2016; Colato *et al*., 2021). Such network changes are thought to be driven by the degradation of structural connections between network regions (Catani and ffytche, 2005; Dineen *et al*., 2009; Schoonheim *et al*., 2015). So far, only one study has shown covarying patterns of WM abnormalities in MS, but several other studies have shown that WM tracts share features which may make them similarly susceptible to pathology (Wahl *et al*., 2010; Li *et al*., 2012, Meijer *et al*., 2016*a*). Understanding the nature of WM degeneration holds the key to elucidating the relationship between functional and structural connectivity and is an important aim in mapping the pathology of cognitive impairment in MS. Thus, in this study we aimed to assess whether WM tracts can be decomposed based on shared pathological or other features, and whether the resulting components reflect known functional network structures.

Our results provide limited evidence of separate covariance structures of WM tracts in MS patients. A single dominant component consisting of all tracts was found in both people with MS and healthy controls, although with somewhat different tract loadings. In MS the main component was largely explained by lesion volume. Even though lesions were masked out of each tract and only non-lesioned tissue was included in the analyses, inflammatory activity in lesions is known to have an effect on surrounding tissue and Wallerian and retrograde degeneration is known to occur in remote areas from the lesions (Trapp *et al*., 1998; Bitsch *et al*., 2000; Trapp and Stys, 2009). After regressing out this predictor, the tract pattern covariance was still shared between all tracts (i.e. no separate patterns of pathology emerged).

A rotation of the first four components (explaining most of the variance in the data) revealed that some tracts loaded more strongly on these components than others. The first component consisted of all the tracts, but those which loaded most highly on this component were large associations tracts connecting distant regions of the brain like the inferior fronto-occipital fasciculus, middle longitudinal fasciculus, optic radiations and vertical occipital fasciculus. Similarly, the second component consisted of association and projection tracts connecting distant brain regions, including the superior thalamic radiation, corticospinal tract and superior longitudinal fasciculus. The third and fourth component consisted of smaller tracts, including commissural tracts, such as the forceps minor and middle cerebellar penduncle. While this suggests differences between types of tracts which will be important to investigate further, these results lack the granularity to draw conclusions about brain networks supported by the tracts in each component. For instance, the first and second components consist of high loadings from tracts which together connect most of the brain. Moreover, most tracts loaded positively on most of the four extracted components, suggesting considerable overlap. This, coupled with the dominant first component prior to rotation, suggests that our results first and foremost show some global aspect of WM microstructure rather than distinct covariance patterns reflecting known functional networks.

There are a number of possible reasons for why pathology patterns associated with functional networks may not have emerged. First, white matter may not show a strong network structure or patterns of covarying pathology. We also found only one dominant tract component in healthy controls, and thus no evidence of a network structure in the white matter (see Appendix 1). Studies which do report patterns of WM pathology have grouped tracts into classes manually, rather than statistically based on shared features (Li *et al*., 2012, Meijer *et al*., 2016*a*). However, despite manual grouping, each class determined by Meijer *et al*., (2016*a*) did show that both FA values and component loadings within a class were associated with cognition, suggesting possible shared damage within a class. Second, patterns of WM pathology may only emerge at later stages of the disease. While network changes measured by rs-fMRI are common in RRMS and occur even in clinically isolated syndrome (CIS, reviewed in Jandric *et al*., 2021), they have been shown to be more pronounced in progressive MS (Meijer *et al*., 2018; Rocca *et al*., 2018). It is therefore feasible that if there is shared susceptibility to MS pathology in the WM, like in the grey matter, it comes more pronounced as the disease advances. This would need to be tested in longitudinal studies or large cross-sectional studies with both RRMS and SPMS samples. Third, patterns of pathology may only become apparent when looking at regions within tracts and not, as assessed in this study, across whole tracts. It is known that many major tracts support several separate functions, for example the interior fronto-occipital fasciculus is involved in cognition and sensorimotor functions as well as other behaviours (Sarubbo *et al*., 2013). Indeed, Li *et al*., (2012) found different FA covariance patterns for different segments of the corpus callosum. In support of this point, a recent study found that WM tract metrics of volume and microstructural integrity from specific section of specific tracts, including subsections of the corpus callosum, superior longitudinal fasciculus and the striato-prefrontal and striato-parietal pathways, better predict cognitive test performance than global tractography and lesion measures, and also better than whole tract measures (Winter *et al*., 2021). This lack of granularity in our data may therefore account for the weak component structure that emerged after rotation. Further studies comparing regional ICA and PCA approaches can help to determine the extent to which each of these factors is at play.

### 4.3 Relationships between WM microstructure and cognition

We found shared variance in WM microstructure across all tracts – this was present in people with MS, despite the heterogeneity of the disease, and determined links to cognitive performance. A large body of literature has now demonstrated associations between WM microstructure metrics and cognitive test performance in MS (Dineen *et al*., 2009; Inglese and Bester, 2010; Hulst *et al*., 2013; Sbardella *et al*., 2013; Llufriu *et al*., 2014, Meijer *et al*., 2016*b*). We identified four cognitive domains: verbal cognition, visuospatial cognition, information processing speed and executive function, consistent with the known domain structure of the BRB-N (Sepulcre *et al*., 2006; De Meo *et al*., 2021).

We found that the first and main tract component was related to specific cognitive domains, but overall these associations were weak. This component, together with sex, lesion volume and normalised brain volume, explained less than 20% of the variance of the verbal cognitive domain. This tract component is made up of most of the tracts investigated, but those which load most highly are long association tracts which connect most of the brain. Interesting, tracts which connect the occipital cortex to the rest of the brain load highly onto this tract component. It may seem as an unexpected finding that tracts associated with visual function predict a cognitive domain without a visual element, but it’s important to consider that we found all tracts to correlate highly with each other, so this correlation between the tract component and cognition is not specific to visual tracts. Nevertheless, damage to the occipital cortex, including atrophy and functional connectivity abnormalities, (in line with known pathology within the optic nerve, i.e. optic neuritis) is commonly reported in MS (Pagani *et al*., 2005; Calabrese *et al*., 2007; Tona *et al*., 2014), so the present finding may reflect a non-cognitively specific marker of MS pathology, albeit weakly, as the tract component only explained a small proportion of this cognitive domain. Lesion volume and atrophy measures were also weak predictors of cognitive domain variance, confirming the clinical-radiological paradox and the need for more advanced brain pathology measures in the study of cognitive impairment in MS.

The weak relationship between test performance on the different cognitive domains and WM tract components contrasts with previous evidence linking WM microstructure in MS to cognitive function (Dineen *et al*., 2009; Inglese and Bester, 2010; Hulst *et al*., 2013; Sbardella *et al*., 2013; Llufriu *et al*., 2014, Meijer *et al*., 2016*b*). This may be due to our use of whole tract measures. There is evidence to suggest that spatial topography is important for cognitive deficits and that some tracts in particular are involved in supporting cognitive function. Most of the early diffusion studies of cognition in MS report correlations between cognitive performance and diffusion metrics in specific regions of tracts, despite analyses being conducted over a whole brain WM skeleton. Those which are commonly reported across the literature are the corpus callosum, cingulum and forceps major and minor (Dineen *et al*., 2009; Sbardella *et al*., 2013; Llufriu *et al*., 2014). In addition, in another study of the sample investigated in the present study, WM metric differences between cognitively impaired and non-impaired patients were found mainly in the corpus callosum and cingulum (Jandric *et al*., 2021*b*). This possibility of spatial specificity has not been formally established through a meta-analysis to date and is therefore speculative. However, it is supported by recent graph theory studies which have found associations between structural connectome metrics in certain networks and cognitive function rather than across the whole connectome (Llufriu *et al*., 2017, 2019; Koubiyr *et al*., 2019; Has Silemek *et al*., 2020). The third tract component identified in this study had had loadings from sections of the cingulum and forceps minor, yet did not explain a great deal of variance of cognitive domains. However, this tract component also consisted of a large number of other tracts with high loadings, so is non-specific to the cingulum and corpus callosum. Future work should focus on establishing if certain WM tracts in particular, e.g. those connecting key hub regions of the brain, are more important for cognitive function and more susceptible to pathology.

### 4.4 Limitations

Our study is not without limitations. Few previous studies have used data decomposition approaches to WM metrics, and those that have used independent component analysis (ICA), which aims to separate sources of signals (Li *et al*., 2012, Meijer *et al*., 2016*a*). In both studies the WM skeleton fed into the ICA returned components which reflected individual tracts or sub-sections of tracts. Grouping of tracts was in both cases done manually, introducing the risk of bias. In contrast, we used PCA to identify shared variance within orthogonal components. This minimises the risk of bias and may also better reflect normal variation in white matter structure. By comparing dominant components between control and patients we were able to evaluate whether such structures are to be expected. However, the possibility should be considered that tract components are not actually orthogonal, perhaps due to the known multifunctional nature of WM tracts, and independent component analysis would in this case have been a more suitable approach. We could also have used factor analytic techniques, but PCA has been shown to produce very similar results to factor analysis when the communalities of variables investigated are greater than 0.7, which was the case in this study (Guadagnoli and Velicer, 1988). This nascent research area requires further detailed work to determine the optimal analysis strategy to identify patterns of white matter pathology. In doing so they can help to understand whether there are networks that are susceptible to MS disease and how these might change over time.

### 4.5 Conclusions and future directions

In this study we have demonstrated that a single dominant component explains the variation in microstructure of white matter in people with MS and in healthy controls. We demonstrate that in people with MS this may relate to the distal effects of WM lesions. These covarying patterns of WM tract variance showed a weak relationship with cognitive function. The study raises several questions about whether, or not, there is a structure to the pathological changes underlying cognitive impairment in MS. Future research should therefore consider whether the effects of lesions spread between tracts and at what stage this may start to impact upon cognitive function. By doing so we can develop a greater understanding of why spatially heterogeneous damage may lead to similar impairments to affect the lives of people with MS.

## Data Availability

The data described in this manuscript are not available in the public domain, as this would not comply with the institutional ethics approval for the project under which the data were acquired. Direct requests for pseudoanonymized data can be considered, and will be dependent on the agreement on an appropriate inter-departmental data transfer agreement that includes the conditions for sharing and re-use.

