## Supplementary material for "A tractometry investigation of white matter tract network structure and relationships with cognitive function in relapsing-remitting multiple sclerosis"

### Appendix 1

**Results for healthy controls**

A PCA of the four microstructural metrics was conducted based on a significant Bartlett’s test of sphericity χ^2^(6) = 40.72, *p* <0.001). Based on scree plot inspection and eigenvalues >1, only the first principal component, which explained 61% of variance, was extracted. The component loadings were 0.92 for FA, -0.91 for RD, 0.86 for MWF and 0.24 for MTR.

Component scores of this principal component (PC) were calculated for each healthy control and each tract and correlated to determine the suitability of a PCA. However, the correlation matrix was not positive definite, suggesting the presence of linear dependencies, due to the small sample size relative to the number of variables. Smoothing was applied to the matrix, and a PCA was still run, but results must be interpreted with caution. The four components before the bend in the screeplot, with an eigenvalue >1, explained 79% of variance. After a Varimax rotation all tracts still loaded positively on rotated tract component (TC) 1. See Figure 1A for the metric and tract correlation matrices and scree plots and Table 2 for the rotated component tract loadings.

Age (r=-0.30), sex (r=-0.28) and intracranial volume (r=0.46) correlated most highly with RC1 weightings (see Figure 1B). In a multiple linear regression model no demographic or anatomical variables predicted TC1 (R^2^ = 0.16, *F*(5, 21) = 2.02, *p* = 0.12). Full statistics for this model are presented in Table 1.

Because no variables explained variance in TC1, for HC the PCA assessing WM tract covariance pattern was not performed again.

Cognitive test scores correlated moderately to highly and a Bartlett’s test of sphericity (χ^2^(36) = 95.64, *p* <0.001) confirmed the suitability of a PCA. The first four principal components explained 82% of variance and were subjected to a Varimax rotation, after which the component weights for cognitive components (CCs) were as follows: SRT C sum (0.93), SRT L sum (0.91) and SRT delayed (0.88) for CC1; WLG (0.88), SDMT (0.71) and PASAT3 (0.60) for CC2; Spatial1to3 (0.90) and Spatial delayed (0.88) for CC3; and PASAT2 (0.96) for CC4, see Table 3. None of the cognitive components was significantly predicted by regression models containing the four WM tract components, age, sex, education, ICV or NVB, see Table 1.

**Table A.1. Predictors of WM tract covariance and cognitive domains**

| **Model** | **Predictors** | **Model statistics** |
| --- | --- | --- |
| **Unrotated tract component 1** | Age: ß = -0.19 *p* = 0.465  Sex: ß = 0.09, *p* = 0.789  Education: ß = 0.01, *p* = 0.937  ICV: ß = 0.56, *p* = 0.101  NBV: ß = 0.14, *p* = 0.605 | R^2^ = 0.16, *F*(5, 21) = 2.02, *p* = 0.12 |
| **CC1: Verbal cognition** | TC1: ß = 1.25, p = 0.258  TC2: ß = -0.10, p = 0.914  TC3: ß = 0.69, p = 0.286  TC4: ß = -2.03, p = 0.056  *Age: ß = -0.68, p = 0.021*  Sex: ß = -0.74, p = 0.055  Education: ß = -0.08, p = 0.623  ICV: ß = -0.77, p = 0.054  NBV: ß = -0.35, p = 0.239 | R^2^ = 0.34, *F*(9, 17) = 2.50, *p* = 0.050 |
| **CC2: Visuospatial cognition** | TC1: ß = -0.32, p = 0.807  TC2: ß = -2.10, p = 0.077  TC3: ß = 0.32, p = 0.682  TC4: ß = 2.05, p = 0.101  Age: ß = -0.03, p = 0.936  Sex: ß = -0.18, p = 0.676  Education: ß = -0.24, p = 0.248  ICV: ß = -0.51, p = 0.266  NBV: ß = 0.36, p = 0.317 | R^2^ = 0.05, *F*(9, 17) = 1.17, *p* = 0.375 |
| **CC3: Information processing** | TC1: ß = -0.63, p = 0.641  TC2: ß = 0.75, p = 0.526  TC3: ß = -1.05, p = 0.198  TC4: ß = 0.86, p = 0.498  Age: ß = -0.19, p = 0.581  Sex: ß = 0.10, p = 0.833  Education: ß = -0.32, p = 0.142  ICV: ß = 0.15, p = 0.746  NBV: ß = 0.11, p = 0.771 | R^2^ = -0.03, *F*(9, 17) = 0.92, *p* = 0.535 |
| **CC4: Executive function** | TC1: ß = -0.93, p = 0.451  TC2: ß = -0.20, p = 0.849  TC3: ß = 0.48, p = 0.503  TC4: ß = 0.38, p = 0.740  Age: ß = -0.16, p = 0.604  Sex: ß = -0.84, p = 0.053  Education: ß = 0.31, p = 0.119  ICV: ß = -0.35, p = 0.422  NBV: ß = 0.19, p = 0.563 | R^2^ = 0.16, *F*(9, 17) = 1.57, *p* = 0.204 |

Significant predictors are presented in italics. Significance threshold *p* < 0.05 applied unless otherwise indicated. Abbreviations: CC = cognitive component , NBV = normalised brain volumeNWMV = normalised white matter volume TC = tract component, WM = white matter

**Table A.2. Tract loadings on each component derived from the tract PCA, after regressing out significant predictors of tract variance and applying Varimax rotation**

| **TC1** | | **TC2** | | **TC3** | | **TC4** | |
| --- | --- | --- | --- | --- | --- | --- | --- |
| **Tract** | **Loading** | **Tract** | **Loading** | **Tract** | **Loading** | **Tract** | **Loading** |
| slf3_r | 0.82 | cbp_l | 0.86 | cbt_r | 0.88 | vof_l | 0.79 |
| slf2_r | 0.80 | cbd_l | 0.77 | cbt_l | 0.82 | vof_r | 0.78 |
| str_l | 0.79 | cbp_r | 0.76 | mcp | 0.56 | fma | 0.74 |
| af_r | 0.78 | atr_l | 0.75 | uf_r | 0.54 | or_l | 0.70 |
| str_r | 0.77 | ar_l | 0.74 | ac | 0.54 | ilf_l | 0.68 |
| cst_l | 0.76 | atr_r | 0.71 | uf_l | 0.46 | or_r | 0.66 |
| slf2_l | 0.74 | fmi | 0.68 | ar_r | 0.46 | mdlf_r | 0.65 |
| fa_l | 0.71 | cbd_r | 0.68 | cbd_r | 0.43 | mdlf_l | 0.63 |
| slf1_l | 0.70 | ifo_l | 0.55 | slf1_r | 0.38 | ifo_r | 0.60 |
| slf3_l | 0.69 | af_l | 0.52 | str_r | 0.38 | ilf_r | 0.57 |
| af_l | 0.69 | mdlf_l | 0.46 | cst_r | 0.36 | cst_r | 0.54 |
| fa_r | 0.67 | fa_r | 0.46 | vof_l | 0.36 | slf1_r | 0.53 |
| ifo_r | 0.64 | fa_l | 0.46 | cbp_r | 0.34 | ifo_l | 0.50 |
| or_r | 0.62 | slf3_l | 0.45 | ar_l | 0.31 | slf3_r | 0.42 |
| cst_r | 0.57 | slf2_l | 0.43 | slf1_l | 0.31 | uf_r | 0.41 |
| ifo_l | 0.56 | str_l | 0.40 | vof_r | 0.31 | slf3_l | 0.41 |
| slf1_r | 0.56 | slf1_l | 0.40 | mdlf_r | 0.27 | slf2_l | 0.39 |
| or_l | 0.53 | ar_r | 0.36 | fa_l | 0.27 | af_l | 0.38 |
| mdlf_l | 0.51 | ilf_r | 0.35 | slf3_l | 0.23 | atr_l | 0.37 |
| uf_l | 0.51 | or_l | 0.35 | cbd_l | 0.22 | slf2_r | 0.37 |
| uf_r | 0.49 | ifo_r | 0.35 | cst_l | 0.20 | af_r | 0.36 |
| fmi | 0.49 | uf_l | 0.35 | fa_r | 0.19 | atr_r | 0.34 |
| mdlf_r | 0.48 | mdlf_r | 0.30 | ifo_r | 0.19 | cst_l | 0.32 |
| ilf_r | 0.46 | af_r | 0.28 | af_r | 0.18 | uf_l | 0.32 |
| atr_r | 0.44 | vof_r | 0.27 | ilf_l | 0.18 | slf1_l | 0.32 |
| fma | 0.42 | cst_l | 0.27 | atr_l | 0.18 | ac | 0.32 |
| cbd_l | 0.41 | fma | 0.26 | ilf_r | 0.18 | fa_r | 0.31 |
| mcp | 0.41 | cst_r | 0.26 | slf2_l | 0.17 | fmi | 0.31 |
| atr_l | 0.40 | ilf_l | 0.24 | slf2_r | 0.14 | cbd_r | 0.31 |
| cbd_r | 0.39 | or_r | 0.24 | atr_r | 0.12 | ar_r | 0.26 |
| ar_r | 0.31 | slf3_r | 0.23 | mdlf_l | 0.12 | ar_l | 0.22 |
| cbp_r | 0.24 | cbt_l | 0.20 | slf3_r | 0.12 | cbt_r | 0.19 |
| vof_l | 0.20 | mcp | 0.18 | or_r | 0.11 | str_r | 0.18 |
| vof_r | 0.18 | uf_r | 0.16 | af_l | 0.10 | cbd_l | 0.16 |
| ilf_l | 0.16 | cbt_r | 0.15 | str_l | 0.09 | cbp_l | 0.16 |
| ar_l | 0.16 | slf1_r | 0.14 | cbp_l | 0.07 | cbt_l | 0.11 |
| cbt_r | 0.11 | ac | 0.10 | or_l | 0.04 | fa_l | 0.10 |
| cbt_l | 0.10 | slf2_r | 0.10 | ifo_l | 0.04 | str_l | 0.09 |
| ac | 0.08 | str_r | 0.09 | fmi | -0.01 | cbp_r | 0.07 |
| cbp_l | 0.07 | vof_l | 0.02 | fma | -0.15 | mcp | -0.22 |

**Table A.3. Cognitive component weights in healthy controls**

|  | **Cognitive RC1** | **Cognitive RC2** | **Cognitive RC3** | **Cognitive RC4** |
| --- | --- | --- | --- | --- |
| **SRT L sum** | **0.91** | 0.08 | 0.10 | 0.02 |
| **SRT C sum** | **0.93** | 0.09 | 0.11 | 0.03 |
| **SRT delayed** | **0.88** | -0.02 | 0.10 | 0.10 |
| **Spatial1to3** | 0.06 | 0.16 | **0.90** | 0.02 |
| **Spatial delayed** | 0.21 | 0.09 | **0.88** | -0.10 |
| **SDMT** | 0.15 | **0.71** | 0.32 | 0.18 |
| **PASAT3** | 0.08 | **0.60** | -0.32 | 0.53 |
| **PASAT2** | 0.09 | 0.11 | 0.01 | **0.96** |
| **WLG** | -0.02 | **0.88** | 0.12 | -0.00 |

Abbreviations: PASAT2 = paced auditory serial addition test 2 second delay; PASAT3 = paced auditory serial addition test 3 second delay; SDMT = symbol digit modalities test; Spatial1to3 = spatial recall test average score over three trials; Spatial delayed = spatial recall test score at the delayed trial; SRT delayed = serial recall test scores at the delayed trial; SRT L sum = serial recall test long term storage sum of scores; SRT L sum = serial recall test consistent recall sum of scores; WLG = word list generation test.

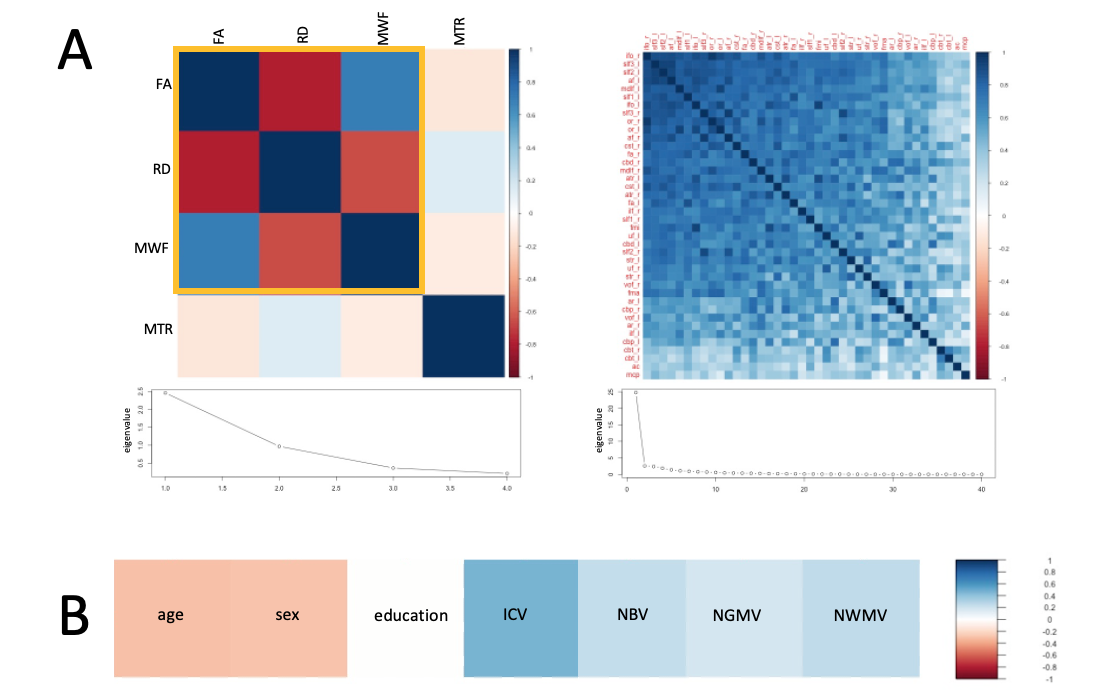

**Figure A.1. Metric and tract principal component analysis in healthy controls**

Figure 1A shows the correlation matrices and scree plots for the PCA ran on the four white matter microstructural metrics (left) and the white matter tracts based on the first component from the metric PCA (right). Those metrics marked with a yellow line load most on principal component 1. All tracts loaded positively on tract principal component 1. Figure 1B shows correlations between rotated tract principal component 1 (TC1) and demographic and anatomical variables. No demographic or anatomical variables predicted TC1 significantly, and the PCA was therefore not repeated, unlike in the MS group.

Abbreviations: FA = fractional anisotropy; RD = radial diffusivity; MWF = myelin water fraction; MTR = magnetisation transfer ratio; ICV = intracranial volume; NBV = normalised brain volume; NGMV = normalised grey matter volume; NWMV = normalised white matter volume
